# Voxel-Wise Quantification and Multi-planar Visualization of Plaque Contrast Enhancement in Intracranial Vessel Wall MRI

**DOI:** 10.1101/2025.10.25.25338801

**Authors:** Yin Guo, Maoxue Wang, Beibei Sun, Dan Cheng, Xin Wang, Kaiyu Zhang, Jiamin Xia, Gador Canton, Duygu Baylam Geleri, Jie Sun, Niranjan Balu, Thomas S. Hatsukami, Mahmud Mossa-Basha, Chun Yuan

## Abstract

**Background:** Contrast enhancement (CE) of intracranial atherosclerotic plaques is a valuable biomarker for identifying culprit plaques in ischemic stroke, but current assessment methods are limited by qualitative approaches and artifact susceptibility.

**Purpose:** To develop and validate a semi-automated method for visualization and quantification of CE intensity and volume, and evaluate its accuracy and scan-rescan reproducibility.

**Study type:** Retrospective.

**Population:** Two patient groups with intracranial atherosclerotic diseases were included: Group A (n=37, 82 plaques) for developing and validating the CE map against expert review, and Group B (n=11, 23 plaques) for assessing scan-rescan reproducibility.

**Field Strength/Sequence:** 3.0 T, 3D time-of-flight gradient echo sequence and T1-weighted fast spin echo sequences, before and after Gadolinium contrast injection.

**Assessment:** A 3D quantitative CE map was developed that incorporated dedicated multi-contrast, multi-planar preprocessing, image intensity normalization, adjustment for enhancement artifacts, and derivation of a signal intensity threshold.

**Statistical Tests:** Receiver operating characteristic curve analysis determined the optimal CE threshold. Sensitivity, specificity, and Cohen’s kappa assessed agreement between the quantitative CE map and manual review. Spearman’s correlation evaluated CE volume quantification. Intraclass correlation coefficient (ICC) and coefficient of variance (CV) assessed reproducibility.

**Results:** The optimal signal intensity threshold for post-contrast maps was 0.4 plus the median plaque wall intensity on pre-contrast maps. The area under the curve for CE detection was 0.91 (95% CI: 0.87-0.94), with sensitivity and specificity of 0.83 and 0.90. CE volume measurements strongly correlated with expert measurements (Spearman’s rho=0.82, p<0.001). Enhancement ratio showed high reproducibility (ICC=0.92, 95% CI: 0.82-0.96), and CE detection demonstrated robust agreement (kappa=0.82, 95% CI: 0.55-1).

**Data Conclusion:** The proposed CE map demonstrated good accuracy compared to expert review and high scan-rescan reproducibility, enabling comprehensive assessment of CE presence, intensity, and volume for evaluating plaque vulnerability in intracranial atherosclerotic disease.

## Introduction

Intracranial atherosclerotic disease (ICAD) is a leading cause of ischemic stroke worldwide(1, 2). Three-dimensional intracranial vessel wall (IVW) magnetic resonance imaging (MRI) has enabled the direct characterization of vessel wall pathologies and is increasingly adopted in both research and clinical settings(3). Among IVW imaging biomarkers, contrast enhancement (CE) of intracranial plaques has emerged as particularly valuable(4–6). Facilitated by gadolinium (Gd)-based contrast agents, plaque CE reflects pathological changes such as increased endothelial permeability and the development of vasa vasorum(7). Recent meta-analyses(8, 9) have highlighted the prevalence of plaque CE in symptomatic ICAD patients, identifying it as the most reliable imaging marker for culprit plaques - plaques most likely to be directly implicated in stroke.

While qualitative approaches have been commonly used in clinical practice(10), recent studies utilizing quantitative analyses of CE on IVW have provided deeper insights, revealing that CE represents a dynamic pathological process. For example, in recently symptomatic ICAD patients(11), CE signals in potentially culprit plaques have been shown to decrease over a one-year period, accompanied by an increase in vessel lumen area. Therefore, quantitative measures of CE signals may offer an unbiased and more precise assessment of clinical risk, while also advancing our understanding of the disease process in ICAD.

A recent study developed a 3D enhancement color map for analyzing intracranial atherosclerotic plaques(12), establishing the genu of the corpus collosum (CC) as a preferred reference structure for image intensity normalization to ensure consistency when comparing plaque CE across patient cohorts. However, additional key capabilities are required to advance quantitative CE analysis. First, the method must effectively model complex, eccentric plaques to enable comprehensive characterization of CE intensity, volume, and other plaque morphological features. Additionally, IVW is susceptible to multiple sources of enhancing artifacts(13), necessitating an adjustment strategy. A visualization framework is also crucial for intuitive depiction of the spatial extent of plaques and their CE characteristics. Finally, reproducible measurement methods for CE are necessary to precisely track subtle changes over time. While the reproducibility of other plaque morphologies has been extensively studied(14, 15), few studies have reported scan-rescan reproducibility for quantitative CE measurements.

Therefore, this study aimed to develop a semiautomatic method for the visualization and quantification of intracranial plaque enhancement, building on a previously proposed multi-contrast, multi-planar framework for ICAD characterization, and to evaluate the accuracy and scan-rescan reproducibility of CE measurements.

## Methods

### Study Design

This retrospective study was conducted in compliance with the Health Insurance Portability and Accountability Act (HIPAA), and the study protocol was approved by the local Institutional Review Board. Written informed consent was obtained from all enrolled subjects, who were prospectively recruited as part of the 3D vessel WALL Imaging (WALLI) study. The study included two independent groups of subjects, all of whom had known intracranial atherosclerotic lesions based on clinical luminal imaging. Group A consisted of 37 patients whose baseline IVW imaging was used to develop the quantitative CE map and establish the intensity threshold for identifying CE(11). Group B consisted of 11 patients who underwent two IVW scans within two weeks(15), to assess the scan-rescan reproducibility of CE measurements using the proposed framework.

### MRI Protocol

All IVW examinations were performed on a 3T whole-body scanner (Ingenia CX, Philips Healthcare, Best, the Netherlands) with a 32-channel head coil. The IVW sequences utilized a 3D Volume ISotropic Turbo spin echo Acquisition (VISTA) protocol with fat suppression and Cartesian UnderSampling with Target Ordering Method (CUSTOM) acceleration, reconstructed using the Self-supporting Tailored k-space Estimation for Parallel imaging (STEP) technique(16). Pre-contrast T1-VISTA (Pre-T1w) and 3D time-of-flight angiography (TOF-MRA) were obtained prior to administering a single-dose gadolinium contrast agent (Prohance, 0.1 mmol/kg at 2cc/s, followed by a bolus of equal volume of saline). Post-contrast T1-VISTA (Post-T1w) was acquired three minutes after contrast injection. MR imaging parameters have been described in previous studies(11, 15).

### IVW preprocessing and plaque annotation

A detailed review protocol of IVW, along with extensive validation of this process, using a multi-contrast, multi-planar framework (MOCHA) has previously been reported(15, 17). Briefly, pre-T1w, post-T1w and TOF-MRA were spatially aligned using rigid image registration, and intracranial artery centerlines were automatically traced on TOF images. One reader identified atherosclerotic lesions on the MOCHA-generated longitudinal and cross-sectional multi-planar reformats (MPRs) across eight arterial segments. The luminal and outer wall boundaries were delineated on the cross-sectional MPRs perpendicular to the luminal centerline. The results were then peer-reviewed by a second reader and disagreements were resolved by consensus. The analyzed arterial segments included the supraclinoid internal carotid arteries (ICA), M1/M2 middle cerebral arteries (MCA), A1/A2 anterior cerebral arteries (ACA), basilar artery (BA), V4 vertebral arteries (VA), and P1/P2 posterior cerebral arteries (PCA).

### Development of the quantitative contrast enhancement map

Building upon MOCHA-based image preprocessing and plaque annotation, IVW images were further processed to generate the quantitative CE map. This process involved the following steps (Figure 1): 1) image intensity normalization, 2) adjustment for enhancement artifacts, including proximity to the sinus and venous flow, 3) deriving the signal intensity threshold for CE, and 4) display and feature calculation.

**Figure 1.**
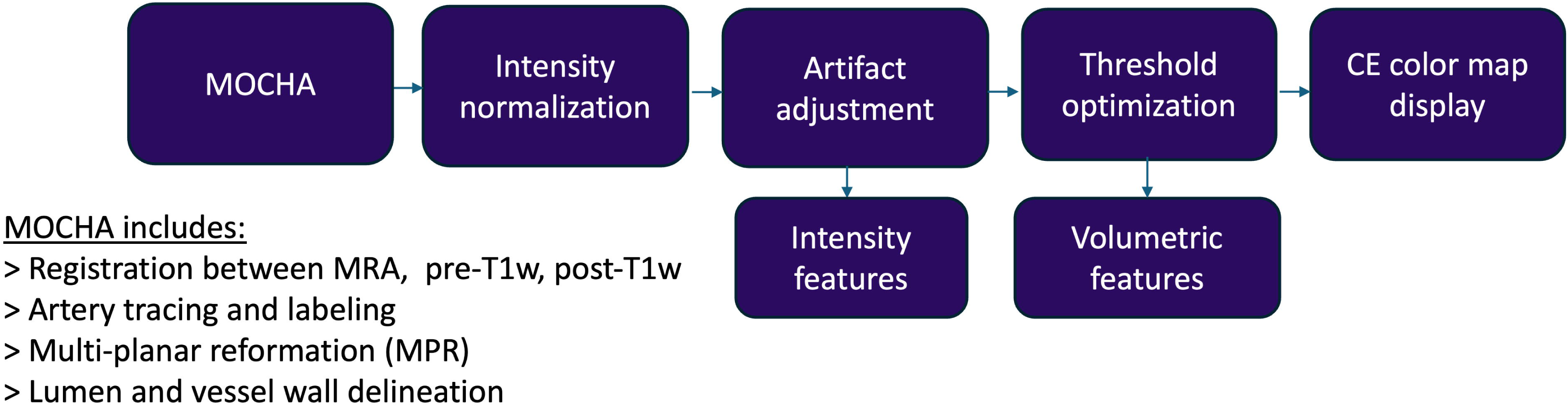
Flow chart of our proposed quantitative contrast enhancement map

#### Image intensity normalization

Following the method described by Omodaka et al.(18), for each subject, a region of interest (ROI) of 20mm^2^ was drawn over a normal appearing region of the genu of the corpus callosum (CC) on the co-registered pre- and post-T1w source images. The average signal intensities (SI) within the ROI were recorded as SI-CC_pre-T1w_ and SI-CC_post-T1w_, respectively. The CC was selected as it is a brain region with uniform signal intensity that is not influenced by contrast enhancement, and previous studies have demonstrated the superiority of CC as a normalizing reference structure compared to other brain regions(19, 20). The normalized pre-T1w and post-T1w maps were then calculated by normalizing each voxel’s intensity to the SI-CC_pre-T1w_ and SI-CC_post-T1w_, respectively, and were applied across all arterial segments within the subject:

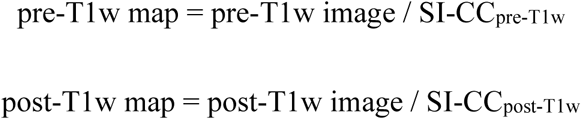

#### Adjustment for enhancement artifacts

Hyperintense signals originating from sinuses and venous flows in post-T1w images were identified as major sources of enhancement artifact(13). Arteries proximal to enhancing sinuses and veins often exhibit elevated CE values, which may not necessarily reflect true vessel wall enhancement, thus compromising the reliability of CE assessment in these regions. To address this issue, a 3D connected components-based algorithm was implemented to identify and isolate CE regions strictly confined within the vessel wall boundary.

Artery regions (R_artery_) were defined by the plaque outer wall boundaries and dilated by 1mm. CE regions (R_CE_) were defined by the 3D connected components where the SI of the post-T1w map exceed 1.0. The CE region within the arterial wall boundary (R_CE-wall_) were derived using an intersection operation:

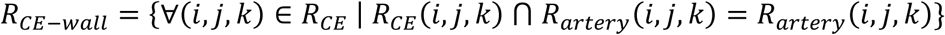

Artifact regions (R_CE-artifact_) were then calculated as:

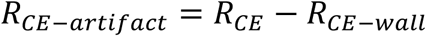

For regions calculated as artifacts (R_CE-artifact_), the post-T1w signal intensity values were adjusted to 0.5. These regions were excluded from subsequent steps of CE detection, segmentation, and feature extraction, as their CE values were deemed unreliable.

Illustrations of the algorithm and examples of common artifacts are shown in Figure 2.

**Figure 2.**
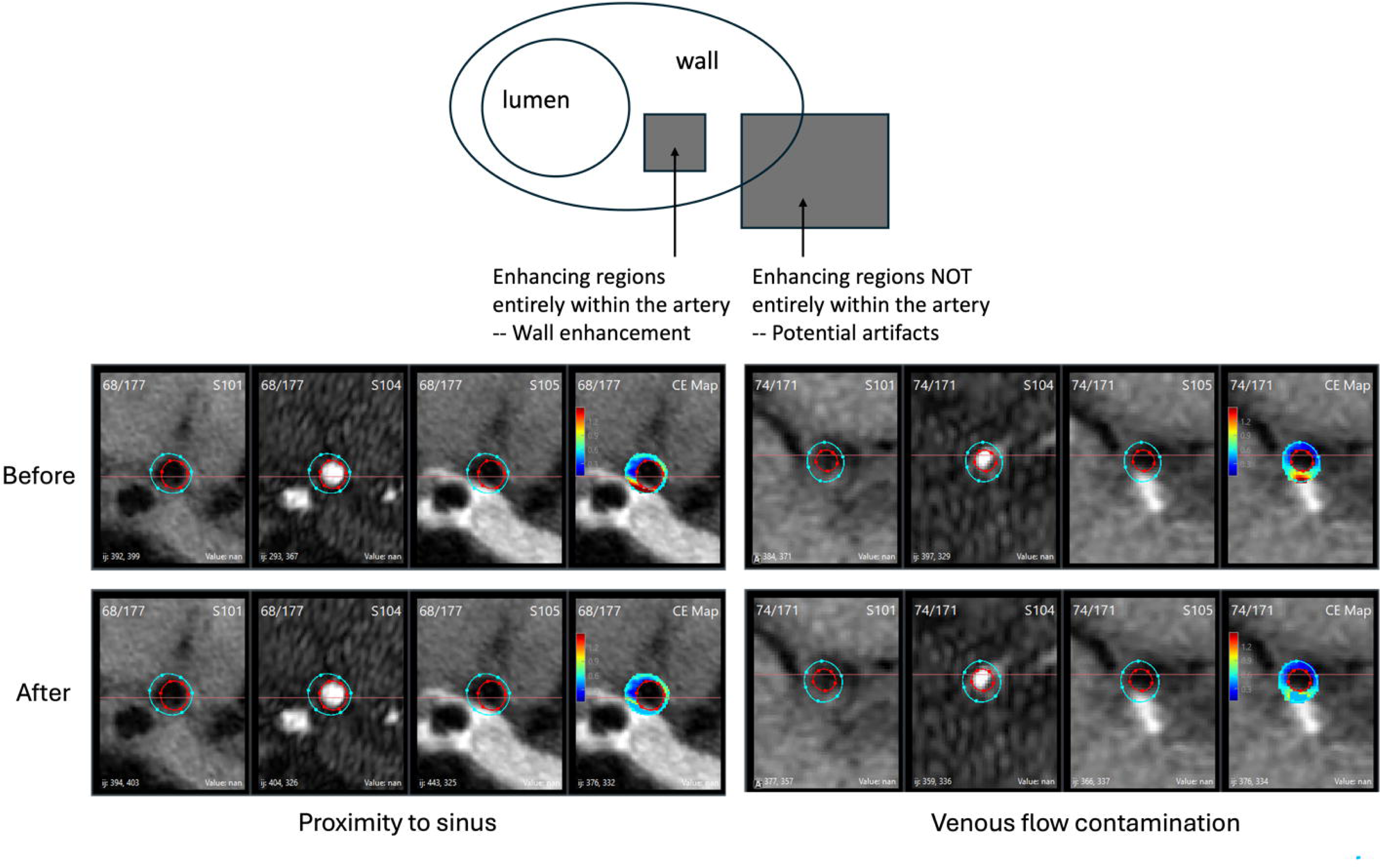
Upper: Illustration of the artifact adjustment process, where a 3D connected component-based algorithm identifies and excludes enhancing regions that extend beyond the arterial wall boundary, reducing false-positive detections from imaging artifacts. Lower: Examples of common artifacts, including proximity to the sinus and venous flow contamination. The proposed algorithm effectively adjusts for these false-positive plaque enhancement signals.

#### Deriving signal intensity threshold for CE

In group A, an expert reader reviewed all identified plaques on the pre- and post-T1w source images, first assessing the presence of CE by visually inspecting for distinct enhancing signals within the artery wall. For plaques with enhancement, the reader classified CE as either grade 1 (enhancement higher than the normal vessel wall but lower than the pituitary infundibulum (PI)) or grade 2 (enhancement similar to or exceeding that of the PI), and further categorized it as focal (localized, well-defined enhancement) or diffuse (widespread, homogeneous enhancement). Additionally, plaque wall regions exhibiting CE were manually segmented using the open-sourced software ITK-SNAP. CE volume within each plaque was then calculated, and the segmentation masks were automatically converted into cross-sectional views in MOCHA. Examples of manual annotation on source images are shown in Figure 3.

**Figure 3.**
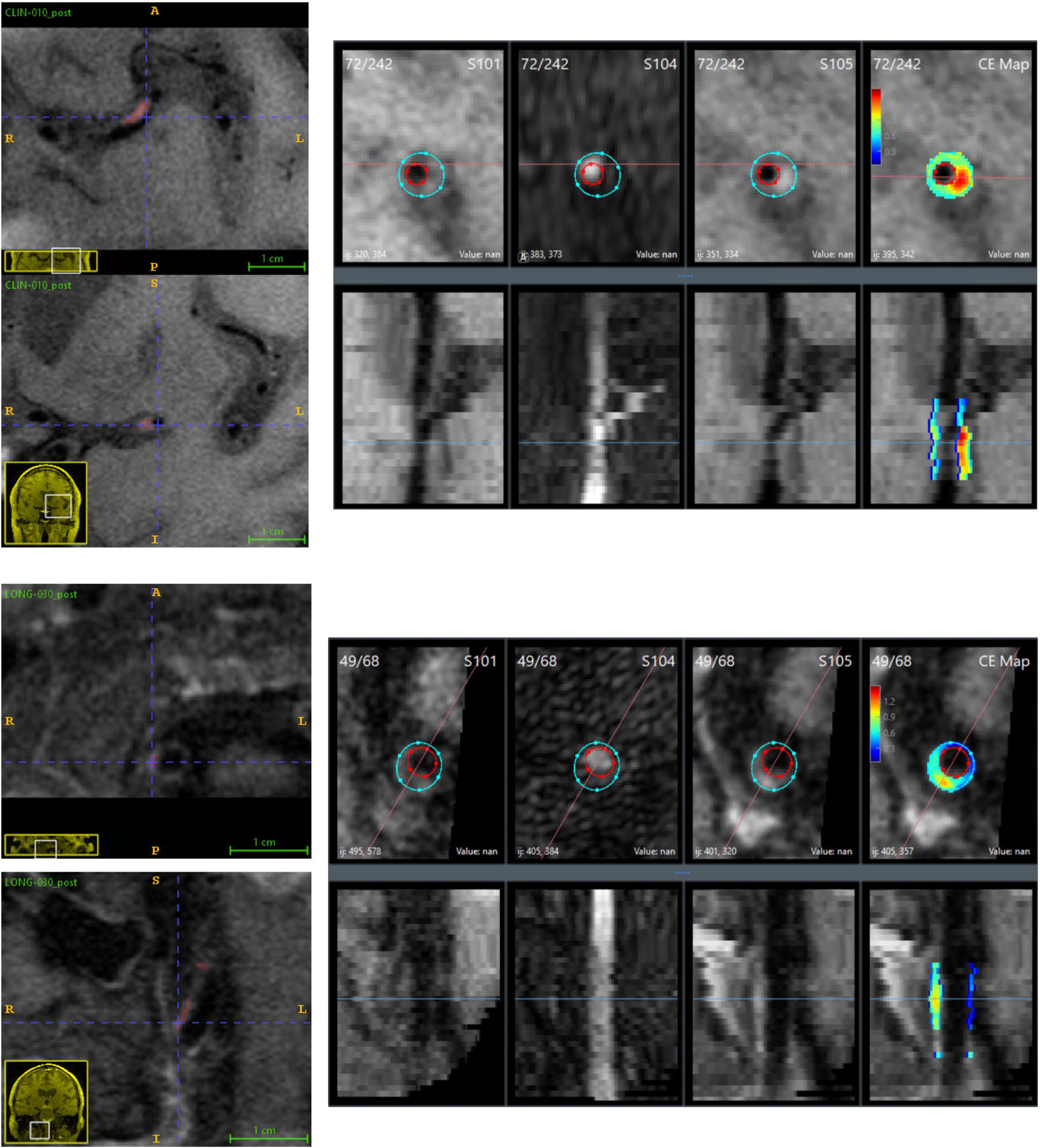
Examples of contrast-enhancing plaques in the upper: left middle cerebral artery and lower: right vertebral artery. The manually segmented enhancement, labeled in the 3D source image (upper: axial view, lower: coronal view), is shown on the left, while the corresponding multiplanar enhancement map visualization is displayed on the right.

To identify the optimal signal intensity threshold for CE, the maximum intensity of the arterial wall from each post-T1 map slice, subtracted by the median intensity of the pre-T1w plaque wall, was compared with the manually determined CE presence or absence (see Statistical Analysis below).

#### Display and feature calculation

The CE color map (within the plaque wall region) was incorporated as an additional contrast weighting in MOCHA’s multi-contrast, multi-planar display, overlaid on the post-T1w images (Figure 3).

We defined the following CE metrics:

1. Mean enhancement ratio: the mean SI on post-T1w map (mean SI on post-T1w images normalized to the CC) within the plaque area The derived signal intensity threshold for CE was applied to all voxels within the plaque area to identify enhancing regions.
2. Gd uptake: the difference between the mean SI on the post-T1w map and the mean SI on the pre-T1w map within the enhancing area.
3. Enhancement volume: the total number of voxels in all enhancing regions, multiplied by the voxel size.
4. Percent enhancement volume: enhancement volume / plaque wall volume * 100%.

### Reproducibility Studies

In group B, an additional image registration step was incorporated into the MOCHA framework to account for the two-week spaced scans. The CC signal intensity was measured separately for each scan. Subsequently, the enhancement metric was calculated automatically.

## Statistical Analysis

In group A, the diagnostic performance of the quantitative CE map was assessed using the area under the curve (AUC) of the receiver operating characteristic curve (ROC). The optimal intensity threshold for CE detection on the normalized post-T1w map was determined by maximizing the Youden index (sensitivity + specificity – 1). Sensitivity and specificity for CE detection using the optimal threshold were estimated at the slice level, with the manual reader annotations as the reference standard.

At the plaque level, the correlation between CE volumes measured on the CE map and manual segmentation was assessed using Spearman’s correlation coefficient, which is less affected by volume changes during MPR processing than measurements of absolute agreement such as the intraclass correlation coefficient (ICC). Additionally, Gd uptake was compared between Grade 1 and Grade 2 enhancement, and CE volume was compared between focal and diffuse enhancement, using generalized estimating equation (GEE)-based regression to account for multiple enhancing plaques within the same subject.

In Group B, scan-rescan reproducibility of CE detection was assessed using Cohen’s kappa. The reproducibility of enhancement ratio, Gd uptake, enhancement volume, and percent enhancement volume was evaluated using the ICC and coefficient of variation (CV).

The nonparametric bootstrap and percentile method was employed to calculate 95% confidence intervals (CIs). Statistical significance was defined as p < 0.05. All statistical analyses were performed using statsmodels 0.13.5 with Python 3.10.

## Results

A total of 37 subjects were included in group A (mean age: 69 ± 12 years, 12 females), and 10 subjects were included in group B (mean age: 69 ± 11 years, 5 females). Clinical characteristics for both groups are presented in Table 1. In total, 82 plaques were identified in the IVW scans of the 37 subjects in Group A, while 23 plaques were detected in both IVW scans of the 10 subjects in Group B.

**Table 1.** Study populations.

| Characteristics | Group A (n = 37) | Group B (n = 10) |
| --- | --- | --- |
| Age (years) | 69 ± 12 | 69 ± 11 |
| Male sex | 25 (68) | 5 (50) |
| Symptomatic | 24 (65) | 0 (0) |
| Hypertension | 29 (78) | 8 (80) |
| Hyperlipidemia | 25 (68) | 5 (50) |
| Diabetes | 9 (24) | 5 (50) |
| Statins use | 29 (78) | 8 (80) |

In group A, there were a total of 673 cross-sectional post-T1w slices from 82 plaques, of which 90 (13%) slices from 17 (21%) plaques were identified as CE-positive by the reader. The overall AUC for CE detection was 0.91 (95% CI: 0.87, 0.94). The signal intensity threshold, determined using the Youden index on the normalized post-T1 map, was derived as 0.4 plus the median intensity of the pre-T1 map (Figure 4a). Applying this threshold, 134 (20%) slices from 23 plaques (28%) were identified as CE-positive. The sensitivity and specificity for CE detection at the slice level were 0.83 and 0.90, respectively.

**Figure 4.**
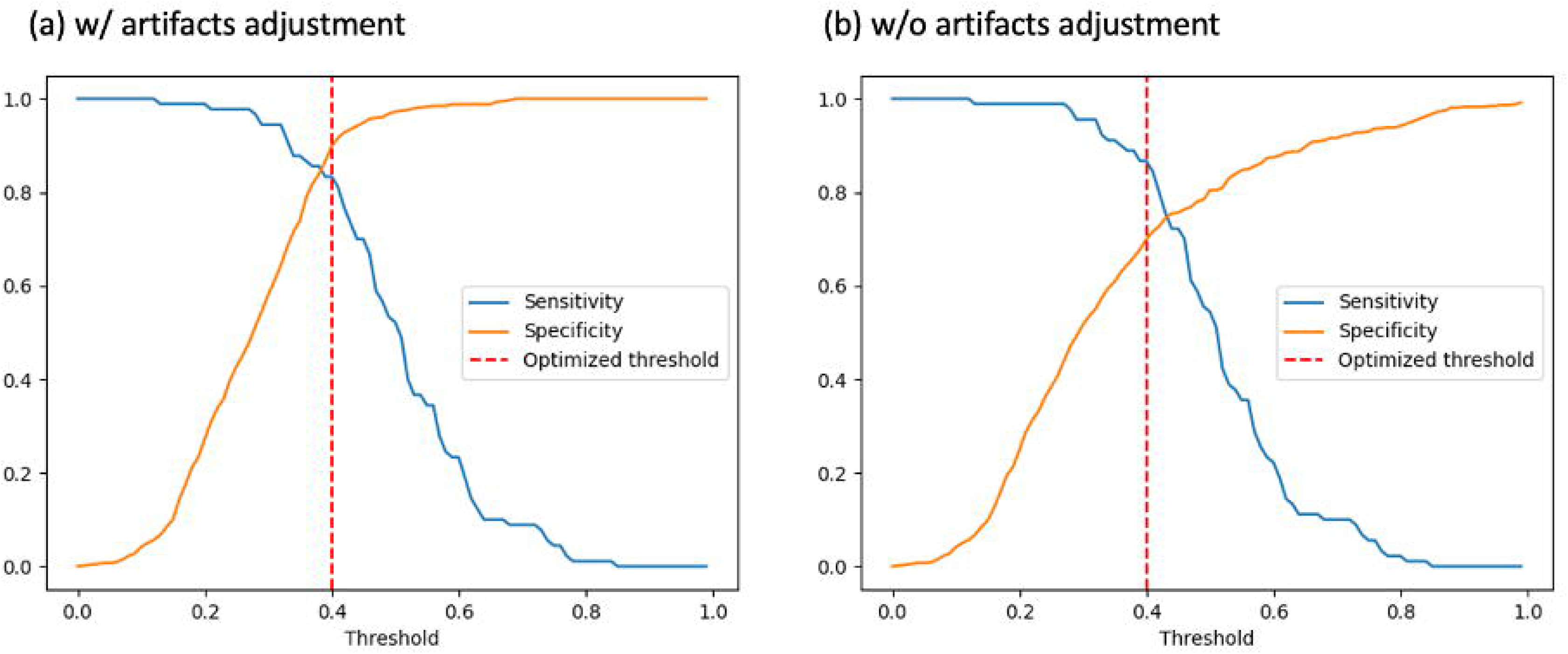
Signal intensity threshold optimization for the presence of contrast enhancement (CE). (a) The optimal signal intensity threshold for CE detection on the post-T1w map was determined as 0.4 plus the median plaque wall intensity of the pre-T1w map, maximizing the sum of sensitivity and specificity. (b) When artifact adjustment was disabled, the same intensity threshold applied; however, a decline in detection specificity was observed.

At the plaque level, 16 plaques were identified as CE-positive by both manual annotation and the quantitative CE map. CE volume measured using the optimal threshold on the CE map strongly correlated with reader measurements (Spearman’s rho = 0.82, p < 0.001). Plaques with Grade 2 enhancement (n = 6) exhibited significantly higher Gd uptake compared to Grade 1 plaques (n = 10, p < 0.001, Figure 5a). Plaques with diffuse enhancement (n = 5) had significantly larger enhancement volume than those with focal enhancement (n = 11, p = 0.03, Figure 5b).

**Figure 5.**
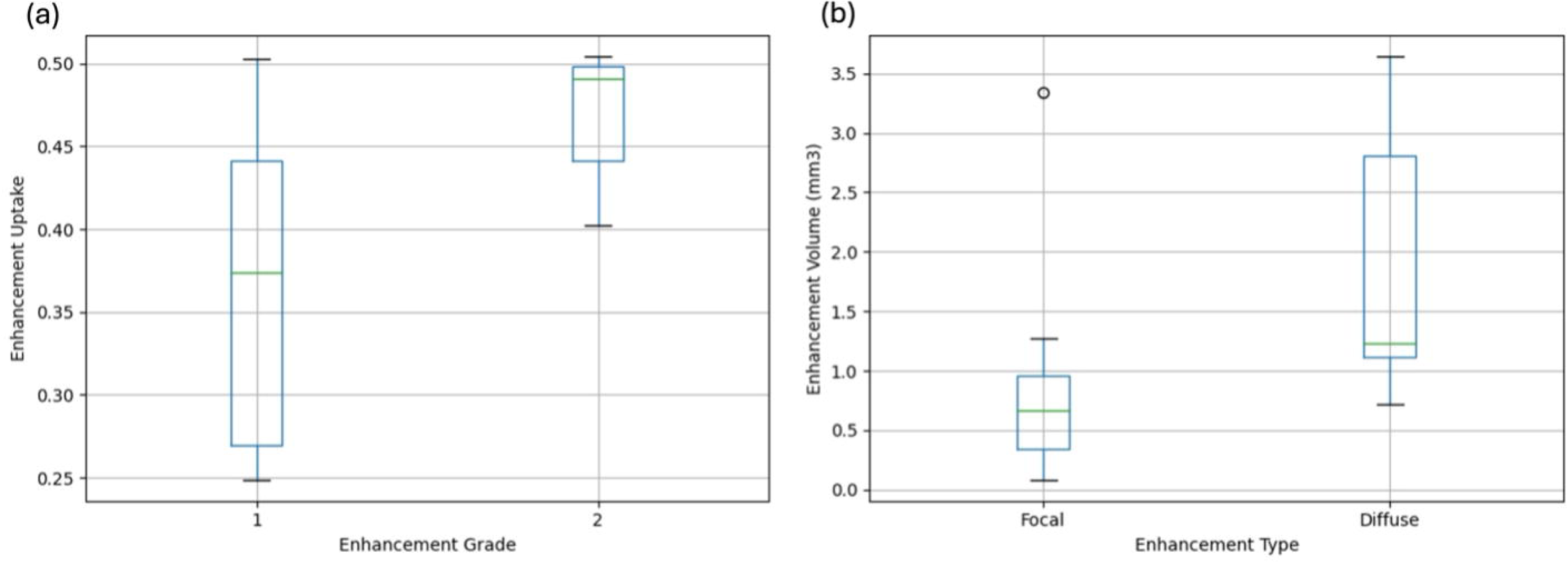
(a) Box plot showing that plaques with Grade 2 enhancement (n = 6) had significantly higher Gd uptake than those with Grade 1 enhancement (n = 10, p < 0.001). (b) Box plot showing that plaques with diffuse enhancement (n = 5) had significantly larger enhancement volumes than those with focal enhancement (n = 11, p = 0.03).

When the modules for artifact adjustment were disabled, with the same threshold (Figure 4b), there was a decline in CE detection performance, with the overall AUC for dropping to 0.78 (95% CI: 0.74, 0.82). Sensitivity and specificity were 0.87 and 0.70, respectively. A total of 253 slices from 39 plaques were identified as enhancing. As signal intensity adjustment only reduces the measured CE values, we observed only a slight decrease in sensitivity after adjustment but a substantial increase in specificity, effectively reducing CE false positives caused by artifacts.

In Group B, the scan-rescan reproducibility of CE signals demonstrated robust agreement. The average histogram overlap (intersection over union, IoU) between the two scans was 0.73 ± 0.12 for pre-T1w maps and 0.75 ± 0.12 for post-T1w maps. The enhancement ratio showed high scan-rescan reproducibility, with an ICC of 0.92 (95% CI: 0.82, 0.96). Examples of scan and rescan CE maps of an internal carotid artery plaque, and their overlapping histograms, are shown in Figure 6.

**Figure 6.**
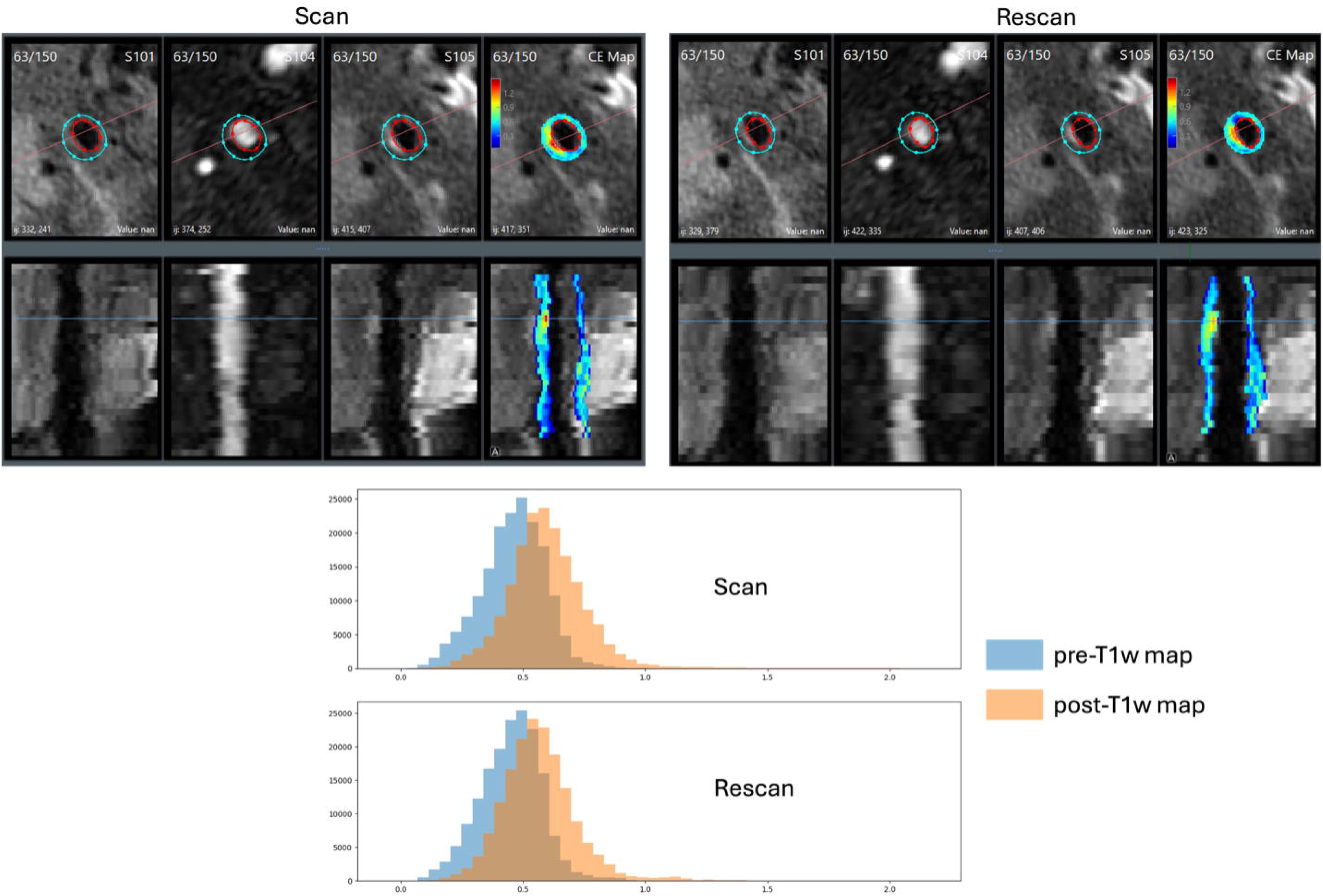
Upper: Example of co-registered scan and rescan intracranial vessel wall images with the corresponding CE map, presented in both cross-sectional and longitudinal views. A supraclinoid internal carotid plaque displaying CE is highlighted. Lower: Histograms of pre-T1w and post-T1w maps demonstrate good overlap between scan and rescan, indicating consistency in normalized signal intensity distributions.

Applying the signal intensity threshold, 9 plaques (39%) were categorized as CE-positive in both scans, 2 plaques (9%) were categorized as CE-positive in only one scan, and 12 plaques (52%) were categorized as CE-negative in both scans, yielding a kappa coefficient of 0.82 (95%CI: 0.55, 1). Among the 9 plaques categorized as CE-positive in both scans, quantitative CE measures showed high reproducibility and small measurement variability (Table 2). The CV statistic favored enhancement ratio, while the ICC statistics favored Gd uptake and volumetric CE measurements. Bland-Altman plots (Figure 7) revealed no apparent relationship between variance and the mean for all the quantitative metrics.

**Figure 7.**
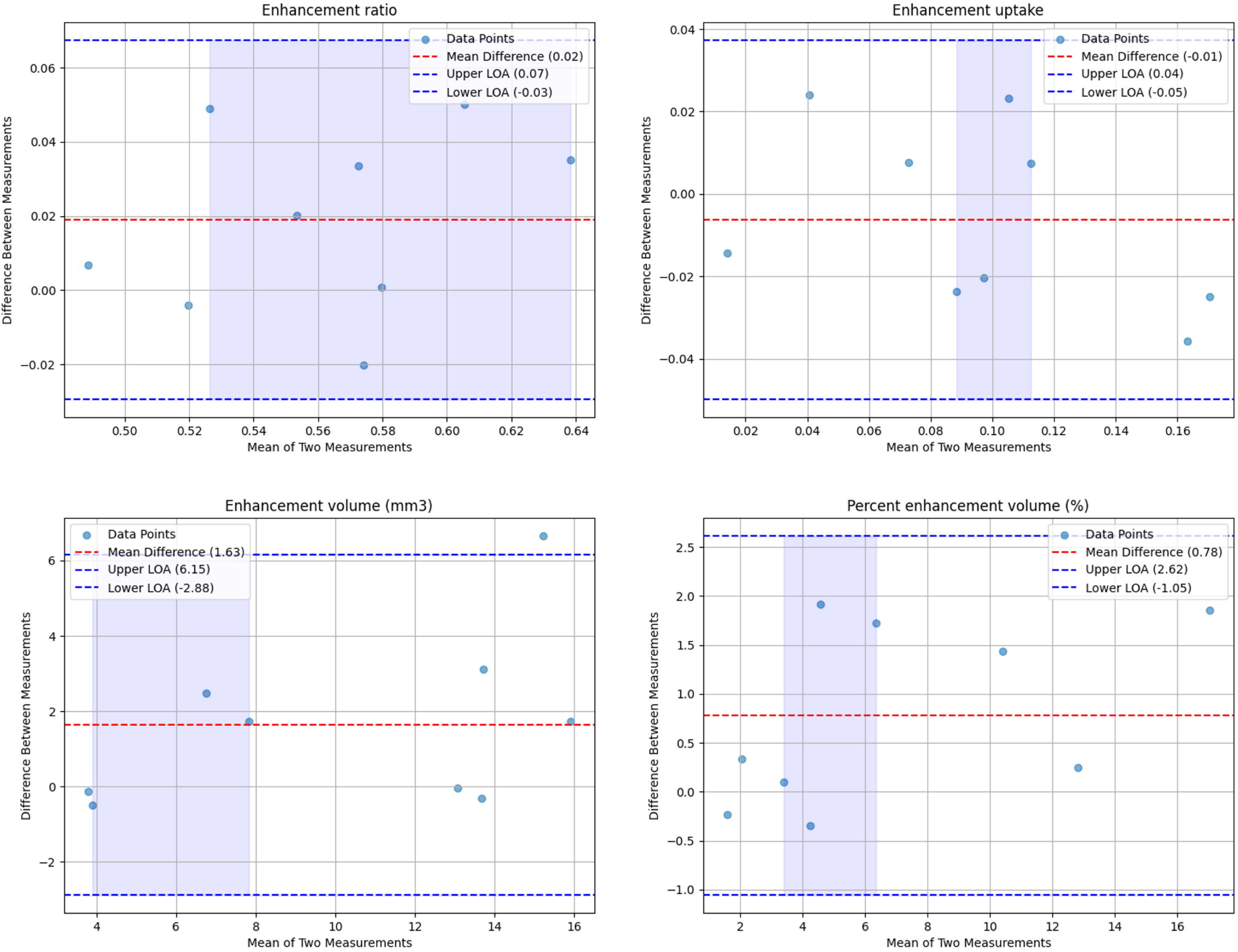
Bland-Altman plots illustrate the reproducibility of enhancement ratio, enhancement uptake, enhancement volume, and percent enhancement volume in the 9 plaques categorized as CE-positive in both scan and rescan sessions.

**Table 2.** Assessment of scan-rescan reproducibility of the quantitative contrast enhancement metrics.

|  | All Plaques (n = 23) |  |  |  | CE-detected plaques in both scans (n = 9) |  |  |  |
| --- | --- | --- | --- | --- | --- | --- | --- | --- |
|  | Scan | Rescan | ICC (95%<br>CI) | CV | Scan | Rescan | ICC (95%<br>CI) | CV |
| Enhancement<br>ratio | $0.55 \pm 0.10$ | $0.54 \pm 0.11$ | 0.92 (0.82,<br>0.96) | 4.4% | $0.57 \pm 0.05$ | $0.55 \pm 0.04$ | 0.81 (0.39,<br>0.95) | 3.0% |
| Enhancement<br>uptake | NA | | | | $0.31 \pm 0.15$ | $0.31 \pm 0.12$ | 0.94 (0.76,<br>0.99) | 10.7% |
| Enhancement<br>volume (mm<sup>3</sup>) | NA | | | | $0.71 \pm 0.34$ | $0.60 \pm 0.28$ | 0.86 (0.51,<br>0.97) | 12.2% |
| Percent<br>enhancement<br>volume (%) | NA | | | | $3.7 \pm 2.8$ | $3.3 \pm 2.6$ | 0.98 (0.90,<br>0.99) | 10.8% |

## Discussion

We developed and validated an image processing method for quantitative characterization of contrast enhancement (CE) in intracranial atherosclerotic plaques, building upon a previously proposed multi-contrast, multi-planar framework for analyzing intracranial vessel wall (IVW) MRI(15, 17). Our method integrated intensity normalization, mitigation of enhancing artifacts, and an optimized signal intensity threshold for CE detection, demonstrating strong agreement with expert reader assessments of CE presence and volume. In addition, the scan-rescan reproducibility of CE detection and measurement was high using the quantitative CE map. Incorporating this method into clinical practice could standardize IVW review and streamline radiologist workflows, enabling comprehensive plaque characterization while improving stroke risk assessment and treatment planning.

Despite the promise of IVW for improving the diagnosis and characterization of intraplaque atherosclerotic diseases, the small caliber of intracranial vessels and the limited resolution of imaging pose significant challenges in identifying specific plaque components, such as intraplaque hemorrhage and necrotic core—features commonly associated with high-risk plaques in larger arterial beds(21–23). Despite these limitations, CE of intracranial plaques has emerged as a valuable biomarker in imaging ICAD. Upon the administration of gadolinium (Gd)-based contrast agents, plaque CE occurs through two primary mechanisms: direct diffusion from the arterial lumen or infiltration via the vasa vasorum in the adventitia. Intriguingly, healthy intracranial arteries lack vasa vasorum(24), which only develops in response to pathological conditions(25). Consequently, the presence of vasa vasorum and associated CE may serve as an indicator of increased endothelial permeability and vessel wall vulnerability. Therefore, CE has been strongly linked to downstream acute infarction(12, 26), with plaques demonstrating CE being up to ten times more likely to cause infarction in their supplied tissue compared to non-enhancing plaques(8). For patients presenting with acute ischemic stroke, integrating CE assessment into routine IVW examinations could serve as a valuable method for identifying high-risk plaques and guiding therapeutic strategies.

Clinicians often assess intracranial plaque enhancement through qualitative grading(26–29) as follows: grade 0 indicates no enhancement; grade 1, enhancement higher than the normal vessel wall but lower than the pituitary infundibulum (PI); and grade 2, enhancement similar to or exceeding that of the PI. Additionally, they qualitatively categorize CE morphology as either focal (localized, well-defined enhancement) or diffuse (widespread, homogeneous enhancement). However, especially in research settings, a broader range of quantitative metrics is desirable for a more detailed analysis of plaque properties. Most studies quantify CE in 2D, using a single image slice from a specific plane of view in the source images, typically selecting the slice with the most severe stenosis or the thickest plaque wall. The degree of CE is then normalized to that of a reference structure, such as the PI(27, 28), gray matter(30, 31), cerebrospinal fluid (CSF)(32), or the genu of the corpus callosum (CC)(12, 33), with the CC shown to provide the most reliable reference for identifying culprit plaques in stroke patients(12).

Despite these advances, most studies have fallen short of achieving intuitive 3D visualizations and voxel-wise CE measurements. Sanchez et al.(12), by adapting a workflow originally designed for quantifying intracranial aneurysms(20, 34), utilized signal intensity probes extending from the lumen into the vessel wall to create a 3D enhancement color map. However, this spoke-based approach often misrepresents eccentric plaques as thin CE surfaces, still requiring manual plaque morphology measurements from source images. Furthermore, a lack of an image registration module between pre- and post-T1w images requires separate measurements on each, doubling the manual efforts for plaques delineation. Our proposed quantitative CE map, built upon MOCHA, addresses these challenges by streamlining ICAD and CE characterization. This approach allows for simultaneous multi-contrast image review, and the organic integration of plaque morphological and enhancement assessments within a single workflow through multi-planar reformation. Beyond MOCHA’s plaque characterization, only the manual signal intensity measurement of the reference structure (CC) is required to generate the CE map, significantly improving efficiency. Moreover, the derived quantitative metrics, including Gd uptake and enhancement volume, were found to correlate well with the widely adopted qualitative metrics.

Additionally, previous quantitative CE maps lacked a definitive threshold for determining CE presence, and these measurements were often compromised by common enhancing artifacts, such as proximity to the sinus or venous flow. While clinicians performing qualitative assessments can often recognize and disregard these artifacts(13), prior quantitative methods have not addressed this issue, leading to false-positive detections and reduced specificity. In our study, we implemented an artifact adjustment module that significantly improved specificity by distinguishing true plaque enhancement from non-plaque-related signal elevations using a 3D connected component-based approach. By excluding enhancing regions extending beyond the arterial wall boundary—likely stemming from imaging artifacts rather than true CE—this adjustment markedly reduced false positives, increasing specificity from 0.70 to 0.90 while maintaining high sensitivity at 0.83. The increased specificity is significant because it ensures that the detected CE truly reflects a pathological process rather than capturing imaging noise, establishing CE as a more reliable IVW imaging biomarker.

The accurate assessment of CE is particularly critical for facilitating longitudinal follow-up studies on ICAD, which may determine the time course of intracranial plaque enhancement and its relationship with acute ischemic events(35, 36). Evidence suggests that both the degree and the persistence of plaque enhancement may serve as promising biomarkers for predicting recurrent strokes(37–39). Given that patients with stroke attributable to ICAD face the highest risk of recurrence among all stroke etiologies(1), this understanding is crucial for optimizing primary and secondary stroke prevention strategies. Several IVW studies have reported different trends in CE changes: plaques associated with recurrent events often exhibit an increase in CE(40), while non-recurrent plaques generally show a decrease in CE(11, 36), potentially indicating a favorable response to medical treatment. To further elucidate the relationship between ICAD and CE evolution, a broader adoption of longitudinal IVW studies is necessary. Our quantitative CE map offers an efficient and reliable method for IVW lesion quantification, which may help overcome the barriers to such future investigations.

Few studies have reported on the reproducibility of CE measurements, which is a critical requirement for reliably tracking changes in plaque enhancement over serial IVW examinations. Existing studies have reported inter-rater ICCs ranging from 0.77 to 0.92(28, 41, 42), although the definitions of the measured quantities vary widely, and these studies continued to rely on traditional 2D approaches to characterize CE intensity. One previous study(15) evaluated the scan-rescan reproducibility of CE signals on IVW, reporting an ICC of 0.94 (95% CI: 0.89, 0.97) for the enhancement ratio in patients scanned twice within a two-week interval. However, that study did not perform intensity normalization, which may be sufficient in an experimental setting, where the same patient is scanned on the same scanner within a short timeframe, but fails to ensure comparability of CE values across patients. This limitation reduces its applicability in larger cohort studies. Importantly, our proposed method was developed and further validated against expert manual review, ensuring reproducibility for broader applicability.

## Limitations

First, while the number of plaques may be sufficient for analysis, this study remains limited by the small number of enhanced plaques. Additionally, scan-rescan validation was performed on a subset of asymptomatic patients, who may exhibit fewer CE features compared to clinical stroke cohorts. Second, the CE threshold was developed and validated against manual review.

Challenges remain in conducting histological studies to establish a definite ground truth for the presence of inflammation in intracranial plaques. Third, this study was conducted at a single center using a single MR scan manufacturer and IVW sequence. In addition, CE intensity may also be influenced by factors related to the contrast injection, such as the timing between gadolinium injection and IVW imaging(32). Future studies are needed to evaluate the generalizability of the CE threshold across different imaging protocols and clinical settings.

## Conclusion

We developed and validated a quantitative CE mapping method for intracranial atherosclerotic plaques, integrating a multi-contrast, multi-planar IVW MRI framework with intensity normalization, artifact mitigation, and an optimized signal intensity threshold for CE detection. Our approach demonstrated high accuracy and strong scan-rescan reproducibility for CE intensity and volumetric measurements. By offering efficient and reliable means of assessing CE, this method has the potential to enhance stroke risk stratification, monitor disease progression, and guide treatment strategies for ICAD patients.

## Data Availability

All data produced in the present study are available upon reasonable request to the authors.

## Notes

### Competing Interest Statement

The authors have declared no competing interest.

### Funding Statement

This work was funded by National Institutes of Health under grants R01-NS092207, R01-NS125635 and R01-NS127317.

### Author Declarations

University of Washington Human Subject Division gave ethical approval for this work.

